# A semi-supervised approach for rapidly creating clinical biomarker phenotypes in the UK Biobank using different primary care EHR and clinical terminology systems

**DOI:** 10.1101/2020.05.14.20101626

**Authors:** Spiros Denaxas, Anoop D. Shah, Bilal A. Mateen, Valerie Kuan, Jennifer K. Quint, Natalie Fitzpatrick, Ana Torralbo, Ghazaleh Fatemifar, Harry Hemingway

**Author notes:** Corresponding author: Spiros Denaxas Institute of Health Informatics University College London 222 Euston Road NW1 2DA, London, UK E. joint second authors (Anoop D. Shah, Bilal A. Mateen).

## Abstract

**Objectives:** The UK Biobank (UKB) is making primary care Electronic Health Records (EHR) for 500,000 participants available for COVID-19-related research. Data are extracted from four sources, recorded using five clinical terminologies and stored in different schemas. The aims of our research were to: a) develop a semi-supervised approach for bootstrapping EHR phenotyping algorithms in UKB EHR, and b) to evaluate our approach by implementing and evaluating phenotypes for 31 common biomarkers.

**Materials and Methods:** We describe an algorithmic approach to phenotyping biomarkers in primary care EHR involving a) bootstrapping definitions using existing phenotypes, b) excluding generic, rare or semantically distant terms, c) forward-mapping terminology terms, d) expert review, and e) data extraction. We evaluated the phenotypes by assessing the ability to reproduce known epidemiological associations with all-cause mortality using Cox proportional hazards models.

**Results:** We created and evaluated phenotyping algorithms for 31 biomarkers many of which are directly related to COVID–19 complications e.g. diabetes, cardiovascular disease, respiratory disease. Our algorithm identified 1651 Read v2 and Clinical Terms Version 3 terms and automatically excluded 1228 terms. Clinical review excluded 103 terms and included 44 terms, resulting in 364 terms for data extraction (sensitivity 0.89, specificity 0.92). We extracted 38,190,682 events and identified 220,978 participants with at least one biomarker measured.

**Discussion and conclusion:** Bootstrapping phenotyping algorithms from similar EHR can potentially address pre-existing methodological concerns that undermine the outputs of biomarker discovery pipelines and provide research-quality phenotyping algorithms.

## Background and Significance

UK Biobank (UKB) is the largest longitudinal research study in the UK (∼500,000 participants), and one of the largest globally [1]. To further enrich this cohort’s data, UKB has begun to link the wealth of information already collected from each individual to their primary care electronic health record (EHR) [1]. In the UK, the first point of contact with the health service for individuals with a new (non-emergency) medical problem or a chronic condition is their local general practitioner (GP). These GPs also receive information from the specialist health services that they refer their patients to, resulting in a closed loop communication system which should result in a complete (time-stamped) summary of their patients’ medical conditions, investigations, regular (prescribed) medications, etc. Introducing primary care EHR information will enable UKB researchers and policy makers to assess the course and outcomes of a plethora of different diseases and risk-factors at scale, whilst allowing them to simultaneously explore the genetic factors associated with each.

Prior to being able to interrogate the data for the approximately 220,000 UKB participants that have already had their data linked, there is the non-trivial task of processing it such that it can meaningfully be interpreted. The primary care data that has been linked to the UKB are derived from the three different countries that compose the UK (England, Scotland and Wales). A total of four data sources (two in England, one in Scotland and one in Wales) using four different controlled clinical terminologies (containing more than 500,000 terms to record information) and different data schemas are used. As a result, researchers with no previous experience working with primary care EHR would need to dedicate a significant amount of time and effort to create phenotyping algorithms for important biomarkers e.g. blood pressure or haematological laboratory markers. This is further complicated by several recent meta-research reports that have identified significant methodological issues with biomarker development research [2], which has resulted in large amounts of research waste [3]. Specifically, these reviews suggest that biomarker discovery pipelines and processes are often plagued by poor methodology [2]; the end result of which is poor replicability of results, as was recently demonstrated by the inability to replicate sample sizes and modelling outcomes in several studies using a large critical care database [4]. Preventing research waste in EHR-based biomarker discovery requires robust clinical validation of phenotypes. However, relying on individual clinical-academics to manually review and refine all of the phenotyping algorithms under development is not easily scalable. As such, an automated but more robust approach for creating and evaluating EHR phenotyping algorithms for biomarkers in primary care data is necessary to address the aforementioned methodological concerns.

One of the primary audiences of this research are US investigators since two-thirds of registered UKB investigators are from US-based institutions. Additionally, the controlled clinical terminologies used in UK EHR data are applicable to US data sources given that CTV3 is a subset of SNOMED-CT. Finally, these challenges are likely not unique to the UK Biobank nor to UK data but exist in other large-scale data resources, e.g. U.S. initiatives such as Electronic Medical Records and Genomics (eMERGE) [5], BioVU [6], Million Veteran Program [7], and All Of Us [8], where primary care EHR data is or will be ingested from multiple disparate data sources and requires significant amount of effort and pre-processing prior to statistical analysis.

The issues of scalability and methodological robustness have become even more relevant in light of UKB announcing that they will be making available the results of COVID–19 tests for participants through Public Health England (PHE), as well as a host of other relevant information, including intensive care data for those who test positive [9]. It is likely that many non-EHR-specialists will be working UKB data for the foreseeable future, and given that the pandemic has already had significant and widespread societal, economic, medical and health service impacts globally [8], there is an impetus to ensure rapid access of these critical data to researchers during this public health emergency, whilst ensuring that this does not come at the cost of research quality [10]. The biomarkers we selected to examine and phenotype in this manuscript are all directly related to modifiable and non-modifiable risk factors for COVID–19 such as diabetes, blood pressure/hypertension, Body Mass Index (BMI) and chronic obstructive pulmonary disease [11–15].

## Aims

The aims of the research presented in this paper are two-fold:

a. To describe a semi-supervised algorithm for rapidly bootstrapping EHR phenotyping algorithms for primary care data by UKB participants;
b. To provide phenotyping algorithms, metadata, implementation details and validation evidence for 31 common biomarkers.

## Methods

### Data sources

#### UK Biobank

Biomarker data for phenotyping were extracted from the UKB, a research study of ∼500,000 adults with extensive phenotypic and genotypic information. Currently, ∼44% of the cohort (n=221,446) have data from primary care EHR linked and made available for researchers (Table 1). Data are collected from English, Scottish and Welsh GP practices that make use of the EMIS (https://www.emishealth.com/), Vision (https://www.visionhealth.co.uk/) or TPP (https://www.tpp-uk.com/) primary care information systems. Data is recorded using four different controlled clinical terminologies: 1) Read version 2 (Read v2); 2) Clinical Terms Version 3 (CTV3); 3) British National Formulary (BNF); and, 4) the Dictionary of Medicines and Devices (DM+D). Both Read v2 and CTV3 are part of the Systematized Nomenclature of Medicine Clinical Terms (SNOMED-CT) [16] and primary care practices in the UK are migrating to using SNOMED-CT terms exclusively.

**Table 1:** **Primary care electronic health record data made available on UK Biobank participants.** EMIS = Egton Medical Information Systems, TPP = The Phoenix Partners, DM+D = Dictionary of Medicines and Devices, BNF = British National Formulary, CTV3 = Clinical Terms Version 3. The number of patients reported was extracted from the registrations table and includes patients with more or one unique registration periods.

| Data Source | Country | Controlled Clinical terminologies: clinical observations | Controlled clinical terminologies: prescriptions | Patients (n) | Clinical events (n) | Prescription events (n) | Data fields |
| --- | --- | --- | --- | --- | --- | --- | --- |
| Vision | England | Read v2 | Read v2 DM+D | 17,860 | 11,973,249 | 6,350,259 | 2 |
| EMIS, Vision | Scotland | Read v2 | BNF | 26,269 | 11,365,300 | 4,301,151 | 3 |
| TPP | England | CTV3 | BNF | 158,894 | 87,493,722 | 39,515,266 | 1 |
| EMIS, Vision | Wales | Read v2 | Read v2 | 20,463 | 12,837,100 | 7,533,324 | 2 |

#### CALIBER

To bootstrap phenotype definitions for the biomarkers of interest we used data from the CALIBER EHR resource. The platform has been described in detail elsewhere [3] but briefly comprises three national EHR data sources: a) longitudinal primary care data from the Clinical Practice Research Datalink (CPRD), b) admitted patient care information on diagnoses and procedures from the Hospital Episode Statistics dataset, and c) cause specific mortality and socioeconomic deprivation information made available from the Office for National Statistics (ONS).

The CALIBER primary care data sourced from the general practices that submit data to the CPRD use the aforementioned Vision software (known as CPRD GOLD), and data are recorded using the Read version 2 clinical terminology system (containing 101,953 terms). In Vision EHR systems, the definition of the data columns is specified by the category of data in the record or the information archetype, which is called an ‘entity type’. For example, the blood pressure entity type specifies that value 1 is the diastolic and value 2 the systolic blood pressure. The associated Read v2 term may contain more details about the subtype of the measurement, e.g. ‘Standing blood pressure reading’.

### Biomarker Data and Unit Recording

In the UKB, measurements from clinical observations (e.g. blood pressure) or laboratory tests (e.g. HbA1c) are recorded with a Read v2 or CTV3 term and up to three structured data columns (value1, value2 and value3). Each data provider uses a varying number of fields to capture information. For example, TPP in England uses a single value, with no explicit or implicit recording of units. Whereas, Scottish data sources are based on three fields, where the second data column (*value 2*) contains the units as free text. Vision-based systems are again different, as the units for recorded values are provided by a separate lookup file and are identified by a unique numeric code. To collate this information, the semi-supervised approach described below captures the relevant unit information across these different structures and processes them into consistent expressions utilizing mapping files to translate inconsistencies to the preferred unit type e.g. *‘x10^9/l’, ‘10^9/L’, ‘x10^9/L’* map to *‘10^9/L’*. Finally, these results were mapped to the Units of Measurement ontology [17](https://www.ebi.ac.uk/ols/ontologies/uo) using a Python script and manually reviewed such that any mismatches could be corrected.

### Semi-supervised phenotyping algorithm bootstrapping

We identified 31 biomarkers spanning blood counts, clinical biochemistry results and physical measurements based on their presumed importance with regards to modelling outcomes for COVID–19 [18], as well as other more generic pathologies such as cardiovascular disease [19], and their availability (recorded at least once during the baseline assessment) (Table 2). In Vision EHR data, groups of similar clinical measurement types or laboratory tests have the same entity type. We used the entity type to identify candidate Read v2 terms which might identify equivalent data items in other data sources. For each biomarker, we performed the following process (Figure 1):

1. We manually mapped UKB fields to Vision entity type identifiers e.g. for lymphocyte counts, the UKB field id is *30210* and the Vision entity type is *208*. We extracted from the UKB showcase information on the units, minimum and maximum value range and mean and identified any relevant unit conversions required.
2. For each Vision entity type, we generated a list of Read v2 terms used to record that biomarker and their frequency. We extracted the Read v2 term with the highest frequency (defined as the “*accepted term”)* e.g. for lymphocytes the term “*42M..00 Lymphocyte count”* was the term most used to record the lab values.
3. We applied a series of automated consistency checks to reduce the number of terms requiring manual review by domain experts. Specifically:

a. We excluded terms that were rarely used i.e. occurring less than 1,000 times and generic Read codes which did not specify the type of biomarker measured e.g. *“4….00 Laboratory procedures”*
b. For lipid measurements, we excluded plasma-based measurements and retained serum derived values. We allowed pre-treatment terms (e.g. pre bronchodilation) but not post-treatment.
c. We excluded terms that did not share the same parent term as the *accepted term* in the Read v2 hierarchy (compared using the first three characters of the Read v2 term) e.g. *“662L.00 24 hr blood pressure monitoring”* was excluded from the blood pressure phenotype where the *accepted term* was “246..00 O/E blood pressure reading”.
4. Using terminology term mappings from the NHS Technology Reference data Update Distribution (TRUD) resource, we mapped non-excluded Read v2 terms to Clinical Terminology Version 3 (CTV3) terms. We only used mappings where the “IS_ASSURED” flag was set to true and included preferred and synonym terms (resulting in some cases to one-to-many maps).
5. We translated the unified list of Read v2 and CTV3 terms into SQL and extracted measurements for all biomarkers across the four data providers iteratively (Supplementary Figure 1).

**Table 2:** **Details on the 31 biomarkers used in this study spanning blood biochemistry, blood count and physical measures.** For units, we provide the UnitOntology entry identifier. The UK Biobank field id column provides the field identifier for the respective biomarker measure, if available, derived from the research data collected at baseline. HDL = high-density lipoprotein, ALP = alkaline phosphatase level, ALP = alanine aminotransferase level, SBP = Systolic blood pressure, DBP = diastolic blood pressure, WBC = White Blood Cell, RBC = red blood cell, CRP = C-reactive protein, MCV = Mean corpuscular volume, MChb conc = Mean corpuscular haemoglobin concentration, FEV1 = Forced Expiratory Volume in 1 second, FVC = Full Vital Capacity

| Phenotype | UK Biobank field id | Phenotype type | units | UnitOntology |
| --- | --- | --- | --- | --- |
| ALP | 30610 | blood biochemistry | U/L | UO_0000179 |
| ALT | 30620 | blood biochemistry | U/L | UO_0000179 |
| Albumin | 30600 | blood biochemistry | g/L | UO_0000175 |
| CRP | 30710 | blood biochemistry | mg/L | UO_0000273 |
| Calcium | 30680 | blood biochemistry | mmol/L | UO_0010003 |
| Cholesterol | 30690 | blood biochemistry | mmol/L | UO_0010003 |
| Creatinine | 30700 | blood biochemistry | umol/L | UO_0010003 |
| Glucose | 30740 | blood biochemistry | mmol/L | UO_0010003 |
| HDL | 30760 | blood biochemistry | mmol/L | UO_0010003 |
| HbA1c | 30750 | blood biochemistry | mmol/mol |  |
| Total bilirubin | 30840 | blood biochemistry | umol/L | UO_0010003 |
| Triglycerides | 30870 | blood biochemistry | mmol/L | UO_0010003 |
| Urea | 30670 | blood biochemistry | mmol/L | UO_0010003 |
| Basophills | 30160 | blood count | 10<sup>9</sup>/L | UO_0000317 |
| Eosinophills | 30150 | blood count | 10<sup>9</sup>/L | UO_0000317 |
| Haematocrit perc | 30030 | blood count | % | UO_0000187 |
| Haemoglobin conc | 30020 | blood count | g/dL | UO_0000208 |
| Lymphocytes | 30120 | blood count | 10<sup>9</sup>/L | UO_0000317 |
| MCHb conc | 30060 | blood count | g/dL | UO_0000208 |
| MCV | 30040 | blood count | fL | UO_0000104 |
| Monocytes | 30130 | blood count | 10<sup>9</sup>/L | UO_0000317 |
| Neutrophills | 30140 | blood count | 10<sup>9</sup>/L | UO_0000317 |
| Platelets | 30080 | blood count | 10<sup>9</sup>/L | UO_0000317 |
| RBC | 30010 | blood count | 10<sup>12</sup>/L | UO_0000317 |
| WBC | 30000 | blood count | 10<sup>9</sup>/L | UO_0000317 |
| DBP | 94 | physical measures | mmHg | UO_0000272 |
| FEV1 | 3063 | physical measures | L | UO_0000099 |
| FVC | 3062 | physical measures | L | UO_0000099 |
| Height | 50 | physical measures | cm | UO_0000015 |
| SBP | 93 | physical measures | mmHg | UO_0000272 |
| Weight | 21002 | physical measures | Kg | UO_0000009 |

**Table 3:** **Descriptive statistics (median and IQR) on the clinical biomarkers defined in this study covering blood counts, key biochemistry markers and physical measurements.** Statistics were stratified by data provider: 1 = England Vision, 2 = Scotland EMIS and Vision, 3 = England TPP and 4 = Wales. HDL = high-density lipoprotein, ALP = alkaline phosphatase level, ALP = alanine aminotransferase level, SBP = Systolic blood pressure, DBP = diastolic blood pressure, WBC = White Blood Cell, RBC = red blood cell, CRP = C-reactive protein, MCV = Mean corpuscular volume, MChb conc = Mean corpuscular haemoglobin concentration, FEV1 = Forced Expiratory Volume in 1 second, FVC = Full Vital Capacity

| Phenotype | Category | UKB id | Units | Eng. Vision events | Eng. Vision median (IQR) | Scotland events | Scotland median (IQR) | Eng. TPP events | Eng. TPP median (IQR) | Wales events | Wales median (IQR) |
| --- | --- | --- | --- | --- | --- | --- | --- | --- | --- | --- | --- |
| ALP | blood biochemistry | 30610 | U/L | 147431 | 73.00 (27.00) | 65017 | 74.00 (28.00) | 907659 | 74.00 (31.00) | 169068 | 73.00 (27.00) |
| ALT | blood biochemistry | 30620 | U/L | 114591 | 23.00 (14.00) | 54724 | 20.00 (12.00) | 891694 | 23.00 (13.00) | 92361 | 22.00 (13.00) |
| Albumin | blood biochemistry | 30600 | g/L | 144475 | 42.00 (4.00) | 33063 | 43.00 (5.00) | 954748 | 42.00 (5.00) | 164884 | 42.00 (5.00) |
| CRP | blood biochemistry | 30710 | mg/L | 23549 | 2.00 (3.00) | 2510 | 3.00 (4.00) | 89406 | 4.00 (3.00) | 18104 | 4.00 (3.30) |
| Calcium | blood biochemistry | 30680 | mmol/L | 54796 | 2.34 (0.14) | 7367 | 2.33 (0.14) | 282652 | 2.35 (0.14) | 42470 | 2.35 (0.14) |
| Cholesterol | blood biochemistry | 30690 | mmol/L | 140222 | 5.20 (1.60) | 75828 | 5.00 (1.70) | 978057 | 5.10 (1.70) | 138801 | 5.10 (1.60) |
| Creatinine | blood biochemistry | 30700 | umol/L | 164360 | 79.00 (23.00) | 102491 | 75.00 (22.00) | 1147028 | 81.00 (23.00) | 204486 | 76.00 (21.00) |
| Glucose | blood biochemistry | 30740 | mmol/L | 41394 | 5.10 (0.90) | 17603 | 5.10 (1.10) | 282383 | 5.10 (1.00) | 28830 | 5.20 (1.10) |
| HDL | blood biochemistry | 30760 | mmol/L | 121635 | 1.40 (0.50) | 46453 | 1.40 (0.57) | 764837 | 1.40 (0.52) | 92854 | 1.30 (0.50) |
| HbA1c | blood biochemistry | 30750 | mmol/mol | 33799 | 40.00 (6.00) | 5915 | 44.00 (8.00) | 175995 | 40.00 (7.00) | 21659 | 41.00 (9.00) |
| Total bilirubin | blood biochemistry | 30840 | umol/L | 137775 | 9.00 (5.00) | 64802 | 9.00 (5.00) | 903641 | 10.00 (6.00) | 140003 | 9.00 (5.00) |
| Triglycerides | blood biochemistry | 30870 | mmol/L | 116308 | 1.30 (0.90) | 40272 | 1.40 (0.93) | 717562 | 1.37 (0.90) | 128464 | 1.36 (0.90) |
| Urea | blood biochemistry | 30670 | mmol/L | 148897 | 5.50 (1.80) | 75630 | 5.60 (1.90) | 1022366 | 5.50 (1.90) | 58332 | 5.30 (1.80) |
| Basophills | blood count | 30160 | 10<sup>9</sup>/L | 129939 | 0.02 (0.10) | 53759 | 0.02 (0.05) | 832814 | 0.02 (0.06) | 138889 | 0.03 (0.10) |
| Eosinophills | blood count | 30150 | 10<sup>9</sup>/L | 130840 | 0.20 (0.10) | 52681 | 0.15 (0.12) | 828186 | 0.16 (0.12) | 142025 | 0.20 (0.12) |
| Haematocrit perc | blood count | 30030 | % | 4055 | 41.10 (5.10) | 11 | 45.00 (5.45) | 11971 | 41.70 (5.30) | 11192 | 41.20 (5.00) |
| Haemoglobin conc | blood count | 30020 | g/dL | 78815 | 13.70 (1.80) | 21512 | 13.60 (1.90) | 903286 | 13.70 (1.70) | 92144 | 13.70 (1.80) |
| Lymphocytes | blood count | 30120 | 10<sup>9</sup>/L | 133853 | 1.80 (0.80) | 50032 | 1.79 (0.82) | 842332 | 1.89 (0.80) | 142637 | 1.80 (0.80) |
| MCHb conc | blood count | 30060 | g/dL | 64138 | 33.50 (1.40) | 28062 | 33.50 (1.70) | 607386 | 33.40 (1.40) | 33023 | 33.60 (1.30) |
| MCV | blood count | 30040 | fL | 136768 | 91.00 (6.00) | 54055 | 90.00 (6.30) | 872756 | 90.90 (6.10) | 147161 | 91.00 (6.00) |
| Monocytes | blood count | 30130 | 10<sup>9</sup>/L | 133696 | 0.50 (0.20) | 52727 | 0.50 (0.27) | 839930 | 0.50 (0.21) | 141545 | 0.50 (0.20) |
| Neutrophills | blood count | 30140 | 10<sup>9</sup>/L | 133588 | 3.40 (1.65) | 53374 | 3.57 (1.86) | 845892 | 3.50 (1.79) | 143045 | 3.50 (1.71) |
| Platelets | blood count | 30080 | 10<sup>9</sup>/L | 137935 | 244.00 (84.00) | 55317 | 249.00 (85.00) | 879866 | 250.00 (84.00) | 145591 | 251.00 (83.00) |
| RBC | blood count | 30010 | 10<sup>12</sup>/L | 135140 | 4.50 (0.58) | 54030 | 4.47 (0.63) | 869832 | 4.55 (0.59) | 146016 | 4.52 (0.58) |
| WBC | blood count | 30000 | 10<sup>9</sup>/L | 140068 | 6.10 (2.25) | 55830 | 6.20 (2.50) | 892739 | 6.25 (2.30) | 147751 | 6.20 (2.30) |
| DBP | physical measures | 94 | mmHg | 357987 | 80.00 (14.00) | 393765 | 80.00 (13.00) | 2833375 | 80.00 (15.00) | 417257 | 80.00 (14.00) |
| FEV1 | physical measures | 3063 | L | 6238 | 2.08 (0.99) | 1430 | 1.79 (0.95) | 47847 | 2.03 (1.04) | 6214 | 2.00 (1.04) |
| FVC | physical measures | 3062 | L | 2792 | 2.95 (1.29) | 233 | 2.96 (1.15) | 32277 | 3.00 (1.31) | 3778 | 2.89 (1.26) |
| Height | physical measures | 50 | cm | 144 | 168.00 (13.12) | 63262 | 167.00 (14.00) | 1069 | 166.00 (15.00) | 27449 | 167.64 (14.98) |
| SBP | physical measures | 93 | mmHg | 358071 | 136.00 (22.00) | 212521 | 135.00 (21.00) | 2836175 | 136.00 (22.00) | 418084 | 137.00 (23.00) |
| Weight | physical measures | 21002 | Kg | 142704 | 77.00 (23.50) | 181172 | 77.90 (23.32) | 1137892 | 78.00 (23.50) | 148988 | 80.00 (25.00) |

**Figure 1.**
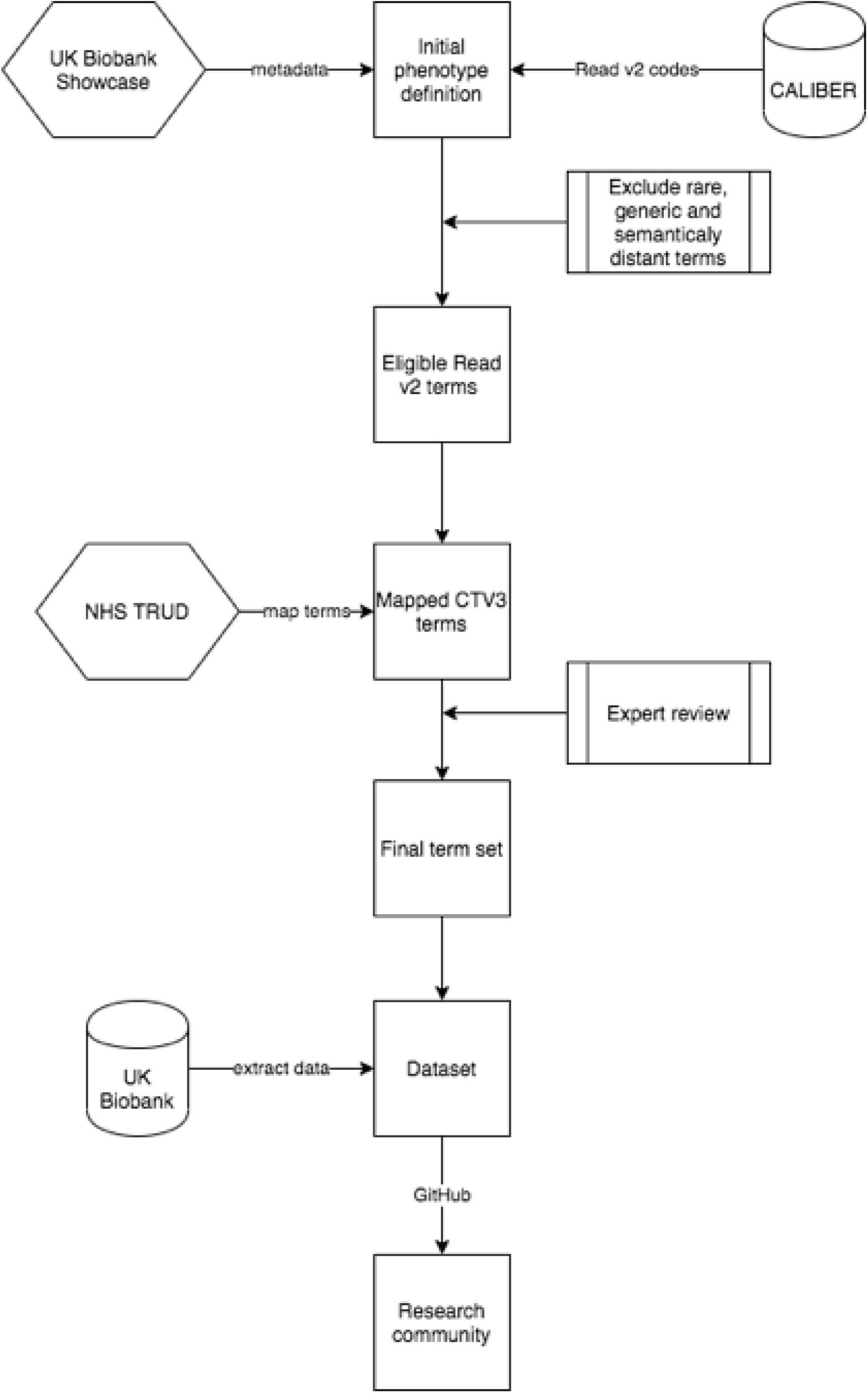
**Description of main steps involved in the semi-supervised approach for rapidly creating electronic health record phenotyping algorithms for biomarkers in the UK Biobank**. The main steps involved in the semi-supervised phenotyping process are: 1) seeding the algorithm definitions using existing phenotype algorithms from the CALIBER resource, 2) excluding generic, rare or semantically distant terms, 3) map Read version 2 terms to Clinical Terms Version 3 terms using the maps provided by the National Health Service (NHS) terminology service (TRUD), 4) expert review and manual inclusion/exclusion of terms, 5) translation to SQL code and data extraction.

### Statistical analyses

We generated and reported descriptive statistics (mean, median, IQR) for extracted biomarker values stratified by provider and plotted the distribution of values using box plots. We compared the distribution of values between data providers for inconsistencies related to data recording. We compared the mean values in the extracted data with the mean biomarker values reported by the UKB baseline values. We calculated and reported sensitivity and specificity per phenotype by calculating Read terms included by the algorithm and subsequently removed upon clinical review, as well as those excluded by the algorithm but which were eventually included following clinical review.

For each biomarker, we fitted a Cox proportional hazards model with all cause mortality as the outcome of interest, adjusted for sex and modelled using restricted cubic splines. We report hazard ratios from the sex adjusted model with 95% confidence intervals.

All analyses were performed using Python v3.7 and the pandas data analysis library (v. 1.0.3, available at https://pandas.pydata.org/). Units were processed using the quantulum3 Python library (v. 0.7.3 available at https://pypi.org/project/quantulum3/).

### Data availability

Unit conversions and mappings, entity type to UKB field mappings, and lists of Read v2 and CTV3 are provided in the Appendix and online https://github.com/spiros/ukb-biomarker-phenotypes The Read v2 to CTV3 mapping file is available from the NHS TRUD service online https://isd.digital.nhs.uk/trud3/user/guest/group/0/home. UKB data can be obtained following approval by applying to the UKB Access Management Committee https://bbams.ndph.ox.ac.uk/ams/. Data in this project was analyzed under protocol ref. 9922 which has been approved by the UKB.

## Results

Using the algorithm described previously, we initially identified 1651 Read v2 and CTV3 terms of which 1,228 were automatically excluded. The majority of terms which are automatically excluded by the algorithm is due to them being marked as “semantically distant” i.e. they do not share a parent term with the most frequently used term for that particular phenotype. Clinical experts reviewed the lists of terms for the phenotypes and manually included terms which were incorrectly flagged for exclusion (false negatives) and conversely removed terms which were incorrectly marked for inclusion (false positives). Specifically, 44 terms were manually included and 103 terms were excluded across all phenotypes. The overall sensitivity of our approach was 0.89 while the overall specificity was 0.92. We calculated sensitivity and specificity estimates for each phenotype and report these in Supplementary Table 2.

This resulted in a final set of 364 unique Read terms were used to extract data. (Figure 2). Using the final set of Read codes, we extracted 38,190,682 events from the GP clinical events table. Of those, 34,578,209 events had a valid measurement attached to them (i.e. not missing and within the valid range). Specifically, we extracted 3,616,003 measurements from England Vision (data provider 1), 1,975,448 measurements from Scotland (data provider 2), 25,233,653 measurements form England TPP (data provider 3) and 3,753, 05 measurements from Wales (data provider 4). Approximately 99.5% of the participants where primary care EHR data were available had at least one biomarker measurement (n=220,981, 52% female).

**Figure 2.**
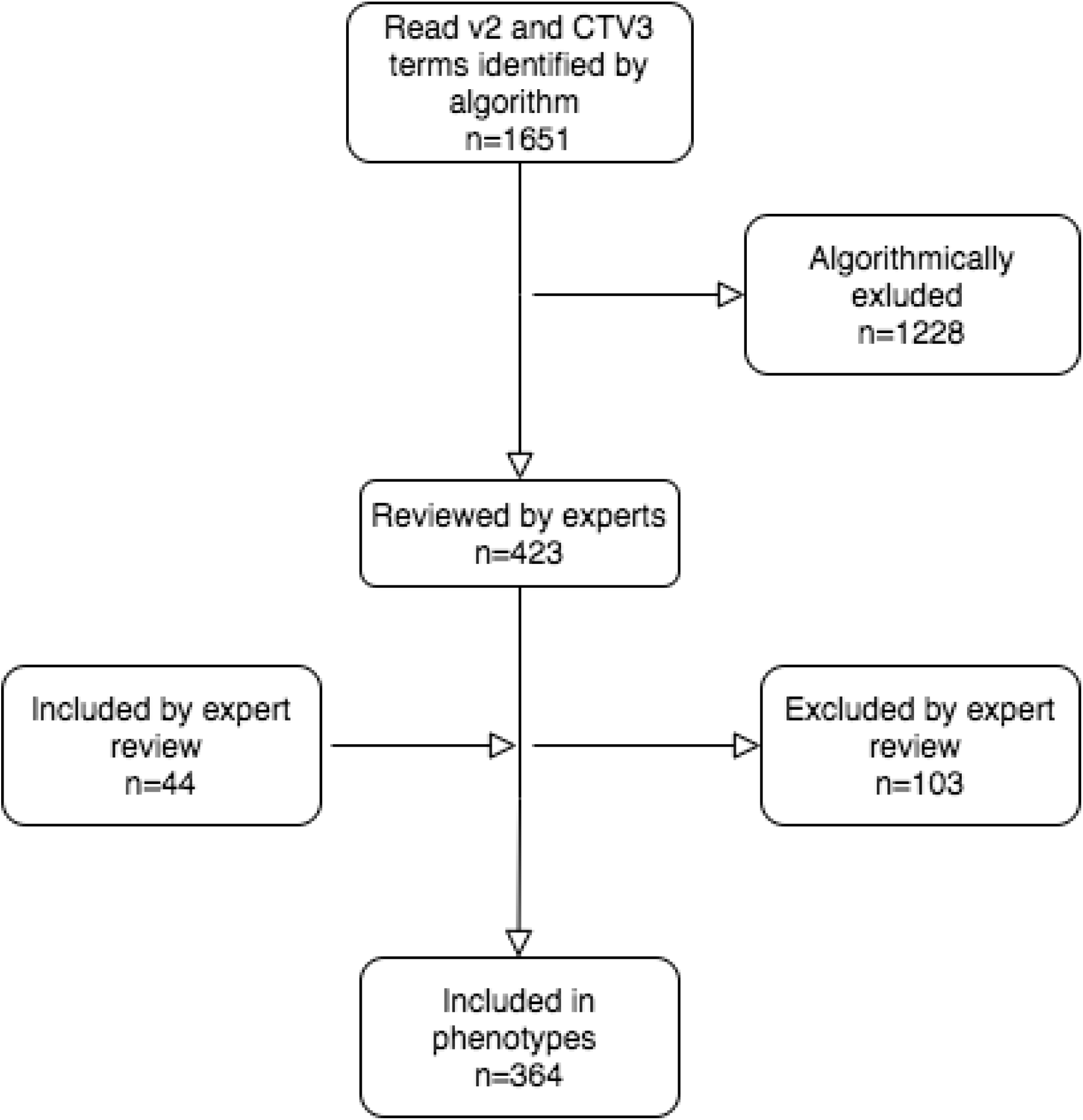
Flow diagram showing the number of Read v2 and CTV3 terms identified by the algorit subsequent inclusions and exclusions performed through expert review. CTV3 = Clinical Terms Version 3

We processed 101 raw unit values recorded in the Scottish data and mapped them to 53 harmonized values. We additionally mapped Vision-specific lookup codes for units to standardized unit definitions (e.g. ME 156 maps to *mmol*). Units were not systematically recorded across the biomarkers with variable levels of missingness: systolic and diastolic blood pressure had units missing in 78% records while FEV1 had units missing in 49%. In contrast, basophils, lymphocytes, monocytes and eosinophils had units recorded for 95% of measurements.

We plotted the distributions of each biomarker across all data sources (Figure 3) and for each data source individually inFigure 4. We calculated and reported descriptive statistics for each biomarker and observed broadly similar distribution values across all sources. Systolic and diastolic blood pressure were the most commonly recorded biomarkers with 3,824,851 and 4,002,384 measurements respectively. The least often recorded marker was Haematocrit percentage with 27,229 values recorded across all data sources.Figure 5. shows Cox proportional hazards regression models using restricted cubic splines and adjusted for sex and age for each biomarker.

**Figure 3.**
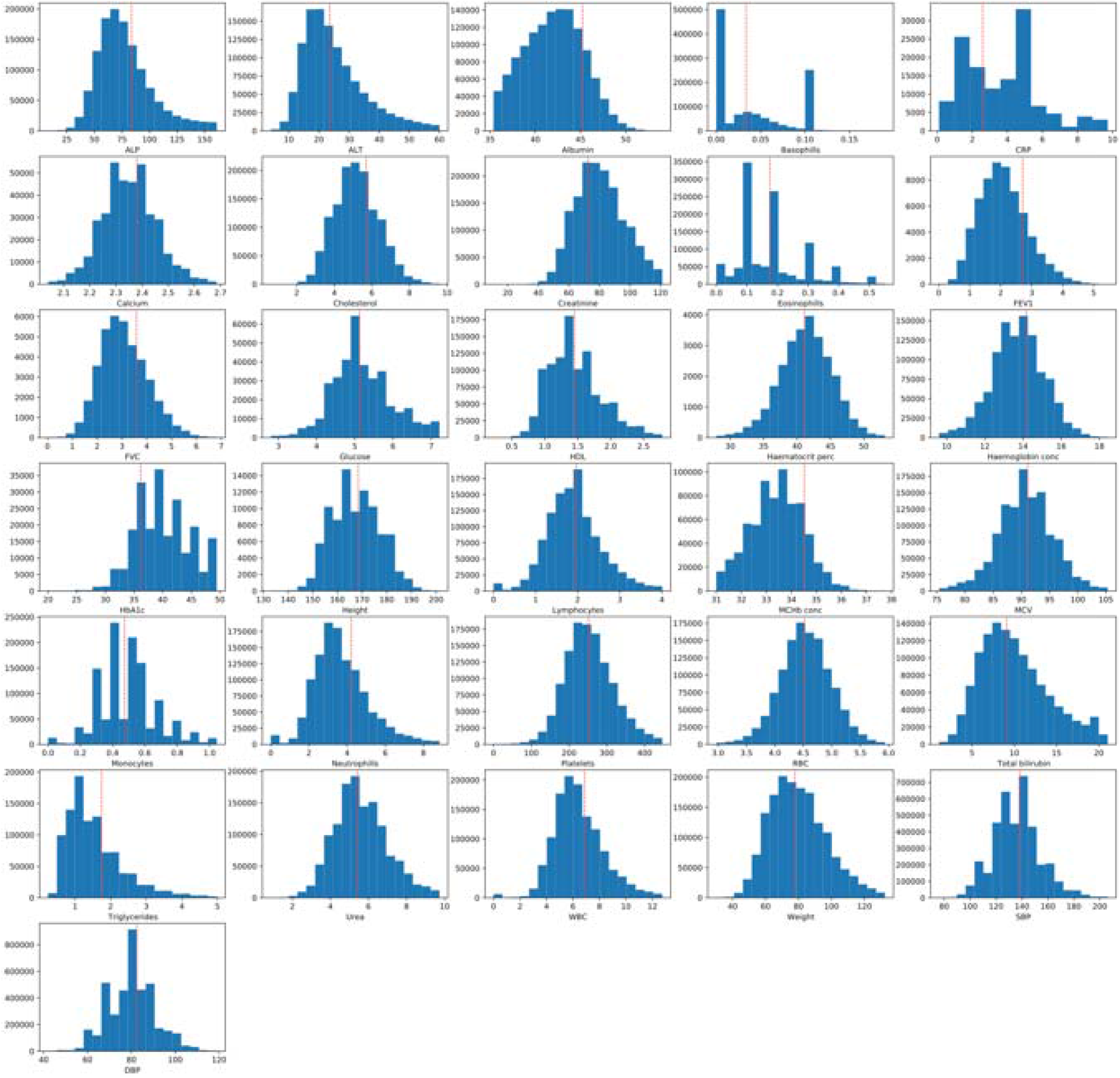
**Histogram plots showing the distribution of values extracted from primary care EHR for the clinical biomarkers defined in this study.** The dashed red line represents the mean value of the bio arker when measured at baseline (across any of the three waves) in study participants (value extracted from the UK Biobank Showcase). Minimum and maximum graph values have been aligned to those reported on the baseline measurements. HDL = high-density lipoprotein, ALP = alkaline phosphatase level, ALP = alanine aminotransferase level, SBP = Systolic blood pressure, DBP = diastolic blood pressure, WBC = White Blood Cell, RBC = red blood cell, CRP = C-reactive protein, MCV = Mean corpuscular volume, MChb conc = Mean corpuscular haemoglobin concentration, FEV1 = Forced Expiratory Volume in 1 second, FVC = Full Vital Capacity.

**Figure 4.**
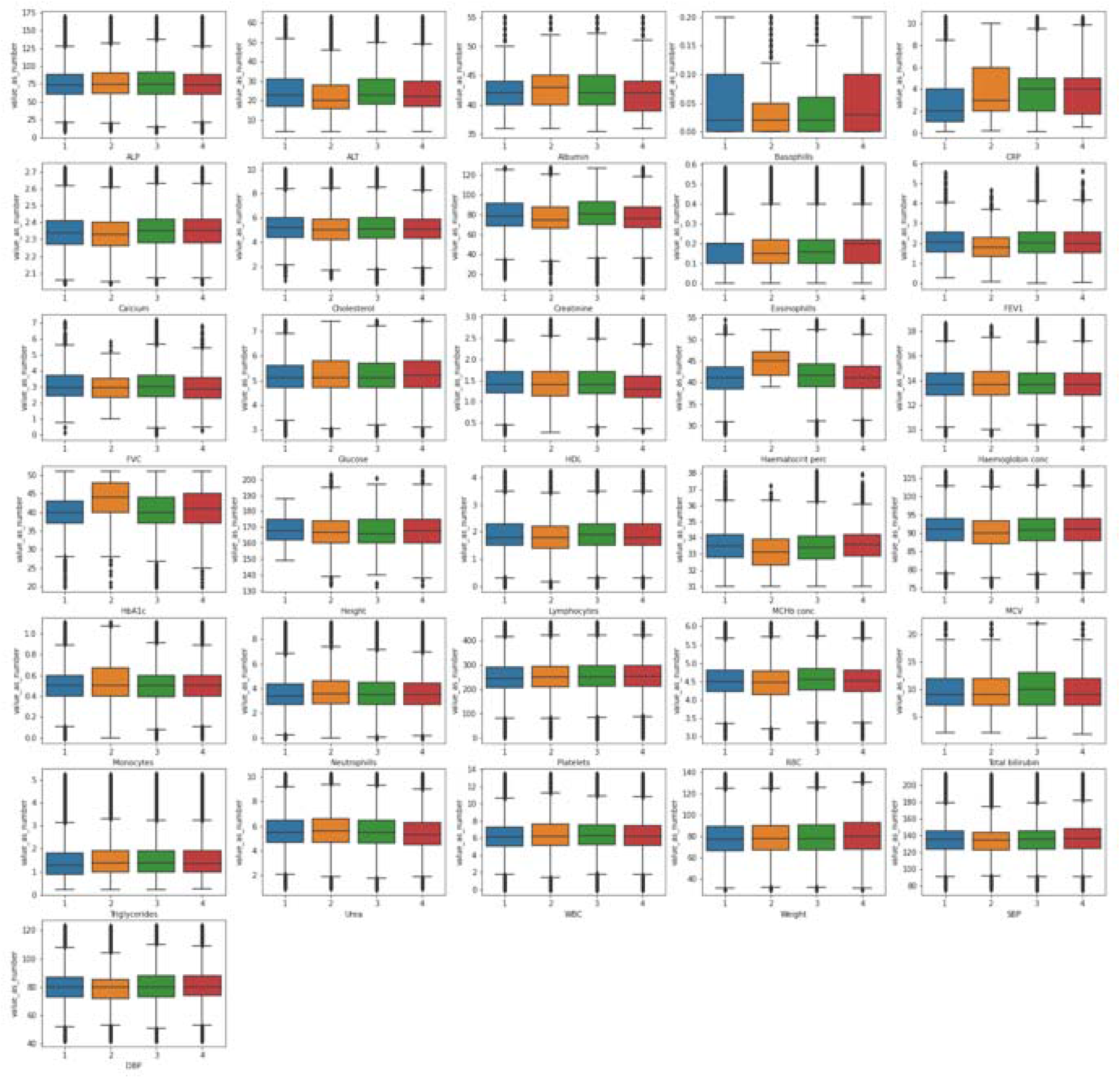
**Boxplot showing the distribution of values extracted from primary care EHR for the clinical biomarkers defined in this study.** 1 = England Vision, 2 = Scotland EMIS and Vision, 3 = England TPP and 4 = Wales. Minimum and maximum graph values have been aligned to those reported on the baseline measurements. HDL = high-density lipoprotein, ALP = alkaline phosphatase level, ALP = alanine aminotransferase level, SBP = Systolic blood pressure, DBP = diastolic blood pressure, WBC = White Blood Cell, RBC = red blood cell, CRP = C-reactive protein, MCV = Mean corpuscular volume, MChb conc = Mean corpuscular haemoglobin concentration, FEV1 = Forced Expiratory Volume in 1 second, FVC = Full Vital Capacity.

**Figure 5.**
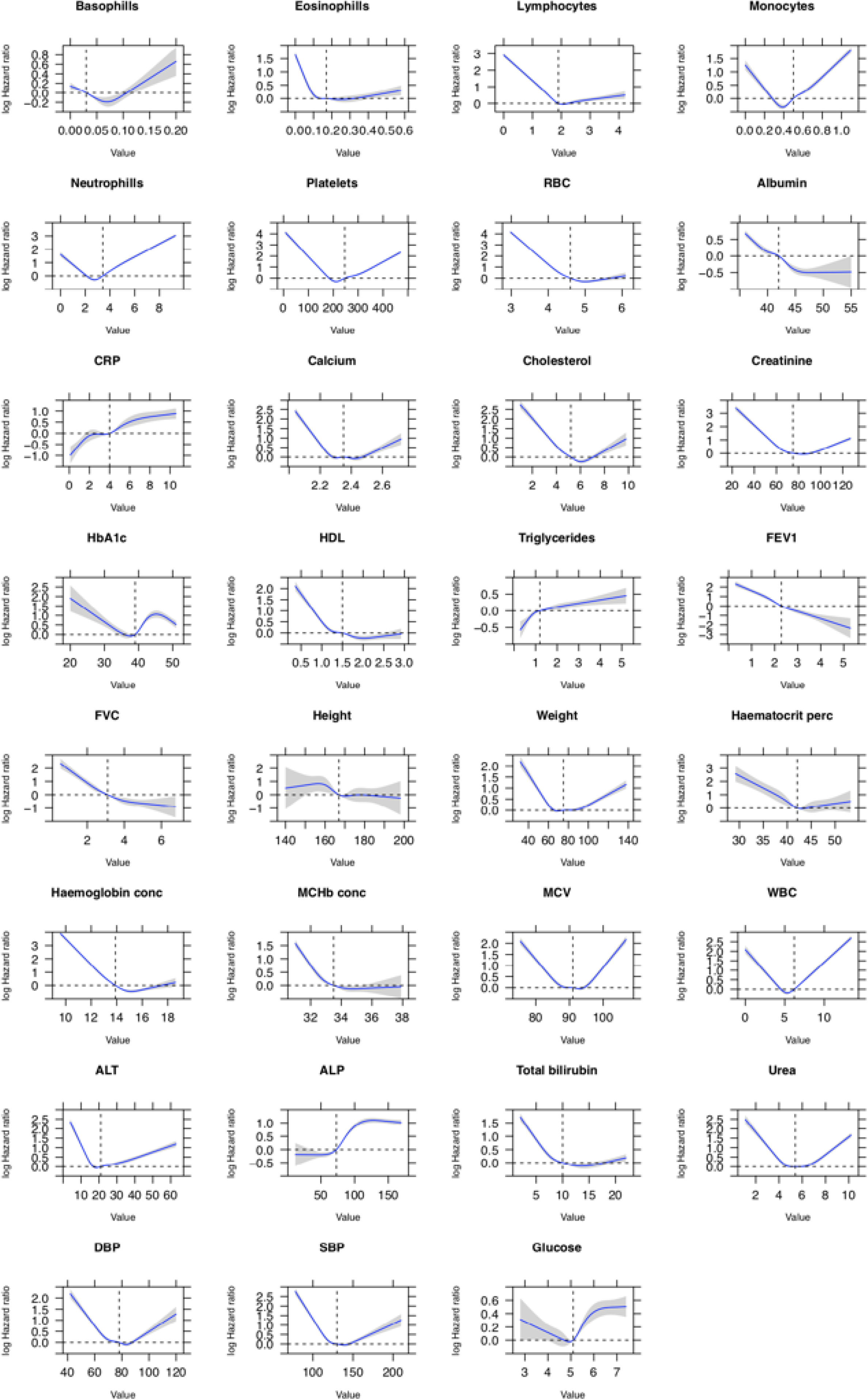
**Adjusted Cox proportional hazards regression restricted cubic spline models for all biomarkers and all-cause mortality. Analyses were adjusted for patient sex and age.** In each panel, the blue line indicates the estimated HR and the grey shading denotes the 95% confidence limits. The horizontal dashed line corresponds to the normal reference hazard ratio of 1.0, values above are associated with increased mortality risk, and values below are associated with decreased mortality risk compared with the reference value. HDL = high-density lipoprotein, ALP = alkaline phosphatase level, ALP = alanine aminotransferase level, SBP = Systolic blood pressure, DBP = diastolic blood pressure, WBC = White Blood Cell, RBC = red blood cell, CRP = C-reactive protein, MCV = Mean corpuscular volume, MChb conc = Mean corpuscular haemoglobin concentration, FEV1 = Forced Expiratory Volume in 1 second, FVC = Full Vital Capacity

## Discussion

In this study, we described a semi-supervised phenotyping approach and applied it on primary care EHR sourced from four different providers in three countries made available for UK Biobank participants. We applied our approach to produce 31 rule-based phenotyping algorithms for commonly used biomarkers. To our knowledge, this is the first study describing how phenotyping algorithms for common biomarkers can be implemented in primary care EHR for UK Biobank participants in a robust and semi-automated manner at scale.

Creating phenotyping algorithms for primary care EHR from four different data sources can be a time-consuming effort requiring a significant amount of effort and resources as they are more than 100,000 potential terms which are inconsistently used to record information. Measurements with different protocols (e.g. BP standing or lying) may be recorded with different Read codes but will have the same entity type. The entity types were used here to give a starting point for Read terms that may be used to record a particular clinical measurement. Additionally, differences in data schemas mean that information is recorded in different ways: for example, data in Scotland can have the units specified as free text in addition to another two values while data from England (TPP) do not specify units and only cover a single value field. Entity types are easier to manipulate given that only a few hundred exist and the group of related Read terms can then be used to identify equivalent terms in CTV3 via the mapping and hence identify equivalent data in the different data sources.

The approach we present here enables researchers to bootstrap algorithms in a robust manner with an overall sensitivity of 0.89 and specificity of 0.92. We observed the lowest sensitivity (0.6) i.e. the highest number of Read terms incorrectly excluded by the algorithm (false negatives) in the glucose phenotype. This was due to the fact that Read v2 terms used to record values did not share a common parent term and were distributed across different branches of the ontology e.g. “*44g..00 Plasma glucose level”* and “*44TA.00 Plasma glucose”*. We observed the lowest specificity estimate (false positives) (0.66) in the red blood cell phenotype where terms related to nucleated red blood cell measurements were included. A similar pattern in terms of specificity was also observed in the Forced Expiratory Volume in 1 second (FEV1) where the initial pool included terms for predicted/expected measurements, post bronchodilation values or terms related to other relevant but not exact measurements (e.g. Forced Vital Capacity FVC).

In line with our phenotyping methodology [20], we evaluated the phenotyping algorithms created by our appoach by estimating hazard ratios adjusted for age and sex with all cause mortality and comparing our findings with known epidemiological associations. We observed similar mortality patterns with previous literature using data extracted from EHR. For example, in line with previous research, we observed an increased risk of mortality in patients with low eosinophil and low lymphocyte counts [21] Similarly, we observed a “U” shape relationship for systolic and diastolic blood pressure measurements which is concordant to previous findings [22]. Finally, intuitively, we observed that a decrease in FEV1 and FVC was associated with an increased risk of mortality and conversely an increase in CRP was associated with an increased risk.

Our method has several strengths. Firstly, it enables the rapid bootstrapping of phenotyping algorithms by reducing the number of Read terms requiring manual review by several orders of magnitude, thereby reducing the amount of resources required. Second, the approach is potentially applicable to non-UK data that face similar challenges, such as for example large biobanked efforts in the US such as MVP and others. Lastly, it provides research-ready phenotyping algorithms for commonly recorded biomarkers in primary care for users. The observed distribution values of the measurements across all biomarkers are consistent with the standard reference ranges for normal results [23]. As previously reported [24] UKB participants are healthier and of higher socioeconomic status than the general population so we would expect to observe these patterns in the measurements.

Our approach also has limitations. Firstly, given that the initial pool of codes is from Read 2 and CTV3 terms are identified through a forward cross-map, it’s possible to omit terms that only exist in CTV3 and are used to record information. The likelihood of this happening however is low given that CTV3 encapsulates Read v2 and GPs tend to use the same set of terms over time. Due to the manner in which codes are used, similar but distinct measurements were sometimes grouped under the same entity code and were incorrectly included by the approach – for example, most lipid measurements had both plasma and serum related terms and manual review subsequently removed the plasma measurements for consistency. Similar patterns were observed, but at much lower frequency, in between fasting and random measurements and values corrected/uncorrected values were reported. Finally, physiological measurements which are performed during routine consultations and not explicitly ordered, such as height, weight and blood pressure required manual phenotyping given the heterogeneity in how data sources captured them despite the small number of Read terms composing the phenotypes (Supplementary Figures 2 and 3).

## Conclusion

In this manuscript, we have demonstrated the challenges that UK Biobank researchers will face when extracting biomarker values from the primary care EHR records of participants. We presented a semi-supervised approach that uses existing phenotyping algorithms and semantic mappings to bootstrap algorithms for 31 common biomarkers spanning haematological and physiological measurements which are widely-used in research. Our research findings are applicable to international audiences given that the controlled clinical terminologies used in the UK primary care EHR are part of SNOMED-CT, two thirds of UK Biobank users US-based investigators and similar large-scale initiatives (e.g. eMERGE, MVP) are likely to face similar challenges. As such, the phenotyping algorithms that have resulted from this work should hopefully facilitate rapid and robust access to the primary care EHR data for UKB participants during the COVID–19 public health emergency, and long after.

## Data Availability

The Read v2 to CTV3 mapping file is available from the NHS TRUD service online https://isd.digital.nhs.uk/trud3/user/guest/group/0/home. UKB data can be obtained following approval by applying to the UKB Access Management Committee https://bbams.ndph.ox.ac.uk/ams/. Data in this project was analyzed under protocol ref. 9922 which has been approved by the UKB.

## Acknowledgements

This research has been conducted using the UK Biobank Resource under Application Number 12345. The views expressed are those of the author(s) and not necessarily those of the National Health Service, the National Institute for Health Research, or the Department of Health. This paper represents independent research [part] funded by the National Institute for Health Research (NIHR) Biomedical Research Centre at University College London Hospital NHS Trust. This study was carried out as part of the CALIBER programme (https://www.ucl.ac.uk/health-informatics/caliber). CALIBER, led from the UCL Institute of Health Informatics, is a research resource consisting of anonymised, coded variables extracted from linked electronic health records, methods and tools, specialised infrastructure, and training and support.

